# Immunogenicity and Safety of a SARS-CoV-2 Inactivated Vaccine (KCONVAC) in Healthy Adults: Two Randomized, Double-blind, and Placebo-controlled Phase 1/2 Clinical Trials

**DOI:** 10.1101/2021.04.07.21253850

**Authors:** Hongxing Pan, Jiankai Liu, Baoying Huang, Guifan Li, Xianyun Chang, Yafei Liu, Wenling Wang, Kai Chu, Jialei Hu, Jingxin Li, Dandan Zhu, Jingliang Wu, Xiaoyu Xu, Li Zhang, Meng Wang, Wenjie Tan, Weijin Huang, Fengcai Zhu

**Affiliations:** NHC Key Laboratory of Biosafety, National Institute for Viral Disease Control and Prevention, Chinese Center for Disease Control and Prevention, Beijing, 102206, China or at; China National Institute for Food and Drug Control, Beijing, 102206, China or at; Jiangsu Provincial Center of Disease Control and Prevention, Nanjing 210009, China or at

## Abstract

**Background:** The significant morbidity and mortality resulted from the infection of a severe acute respiratory syndrome coronavirus 2 (SARS-CoV-2) call for urgent development of effective and safe vaccines. We report the immunogenicity and safety of a SARS-CoV-2 inactivated vaccine, KCONVAC, in healthy adults.

**Methods:** Two phase 1 and phase 2 randomized, double-blind, and placebo-controlled trials of KCONVAC were conducted in Chinese healthy adults aged 18 through 59 years. The phase 1 trial was conducted in a manner of dosage escalation. The first 30 participants were randomized in a ratio of 4:1 to receive two doses of either KCONVAC at 5 μg per dose or placebo on Day 0 and Day 14, and the second 30 participants were randomized to receive either KCONVAC at 10 μg per dose or placebo following the same procedures. The participants in the phase 2 trial were randomized in a ratio of 2:2:1 to receive either KCONVAC at 5 μg or 10 μg per dose, or placebo on Day 0 and Day 14, or Day 0 and Day 28. In the phase 1 trial, the primary safety endpoint was the proportion of participants experiencing adverse reactions/events within 28 days following each vaccination. Antibody response and cellular response were assayed in the phase 1 trial. In the phase 2 trial, the primary immunogenicity endpoint was the seroconversion and titre of neutralization antibody, and the seroconversion of receptor binding domain (RBD)-IgG 28 days after the second dose.

**Findings:** In the phase 1 trial, 60 participants were enrolled and received at least one dose of 5-μg vaccine (N=24), 10-μg vaccine (N=24), or placebo (N=12). In the phase 2 trial, 500 participants were enrolled and received at least one dose of 5-μg vaccine (N=100 for 0/14 or 0/28 regimens), 10-μg vaccine (N=100 for each regimen), or placebo (N=50 for each regimen). In the phase 1 trial, 13 (54%), 11(46%), and 7 (58%) participants reported at least one adverse event (AE), of whom 10 (42%), 6 (25%), and 6 (50%) participants reported at least one vaccination-related AE after receiving 5-μg vaccine, 10-μg vaccine, or placebo, respectively. In the phase 2 trial, 16 (16%), 19 (19%), and 9 (18%) participants reported at least one AE, of whom 13 (13%), 17 (17%), and 6 (12%) participants reported at least one vaccination-related AE after receiving 5-μg vaccine, 10-μg vaccine, or placebo at the regimen of Day 0/14, respectively. Similar results were observed in the three treatment groups of Day 0/28 regimen. All the AEs were grade 1 or 2 in intensity. No AE of grade 3 or more was reported. One SAE (foot fracture) was reported in the phase 1 trial. KCONVAC induced significant antibody response. 87·5% (21/24) to 100% (24/24) of participants in the phase 1 trial and 83·0% (83/100) to 100% (99/99) of participants in the phase 2 trial seroconverted for neutralising antibody to live virus, neutralising antibody to pseudovirus, and RBD-IgG after receiving two doses. Across the treatment groups in the two trials, the geometric mean titres (GMTs) of neutralising antibody to live virus ranged from 29·3 to 49·1 at Day 0/14 regimen and from 100·2 to 131·7 at Day 0/28 regimen, neutralising antibody to pseudovirus ranged from 69·4 to 118·7 at Day 0/14 regimen and from 153·6 to 276·6 at Day 0/28 regimen, and RBD-IgG ranged from 605·3 to 1169·8 at Day 0/14 regimen and from 1496·8 to 2485·5 at Day 0/28 regimen. RBD-IgG subtyping assay showed that a significant part of RBD-IgG was IgG1. The vaccine induced obvious T-cell response with 56·5% (13/23) and 62·5% (15/24) of participants in 5-μg and 10-μg vaccine groups showed positive interferon-γ enzyme-linked immunospot responses 14 days after the second dose in the phase 1 trial, respectively.

**Interpretation:** KCONVAC is well tolerated and able to induce robust antibody response and cellular response in adults aged 18 to 59 years, which warrants further evaluation with this vaccine in the upcoming phase 3 efficacy trial.

**Funding:** Guandong Emergency Program for Prevention and Control of COVID-19 (2020A1111340002) and Shenzhen Key Research Project for Prevention and Control of COVID-19.

## Introduction

The emergent severe acute respiratory syndrome coronavirus 2 (SARS-CoV-2) caused a pandemic of coronavirus disease 2019 (COVID-19) as declared by the World Health Organization (WHO) on March 11, 2020.^1,2^ The virus is highly transmissible. As of February 19, 2021, near 110 million cases and more than 2·4 million deaths have been reported worldwide.^3^ The significant morbidity and mortality call for an urgent need for effective and safe vaccines against COVID-19. As of February 17, 2021, there are 69 SARS-CoV-2 candidate vaccines in various phases of clinical development, and 181 in preclinical development according to WHO.^4^ A variety of platforms or technologies are applied for the development of these vaccine, including inactivated vaccine, adenovirus vectored vaccine, recombinant protein-based vaccine, RNA vaccine, and DNA vaccine. ^4^ The safety, immunogenicity, and/or efficacy in human have been reported for these vaccines, demonstrating good immunogenicity, efficacy, and acceptable safety profile. ^5-15^ The inactivated vaccines against various infectious diseases have been used for decades which confers them some advantages such as well-documented safety record, well developed and matured manufacturing process, able to present multiple viral proteins for immune recognition, among others. Two inactivated SARS-CoV-2 vaccines manufactured by Beijing Institute of Biological Products/Sinopharm and Sinovac have received conditional approval in China. The expected enormous gap between the need for vaccine and manufacturing capability drives development of more vaccines. Here we report the preliminary analysis of immunogenicity and safety from two ongoing phase 1/2 clinical trials with an inactivated SARS-CoV-2 vaccine, called KCONVAC, which were conducted in Chinese adults.

## Methods

### Study design and participants

Both the phase 1 and phase 2 trials of KCONVAC were randomized, double-blind, and placebo-controlled studies and conducted in succession by Jiangsu Provincial Center for Disease Control and Prevention (JPCDC) beginning from Oct 2020. The studies were done in accordance with the Declaration of Helsinki and Good Clinical Practice. An independent data safety monitoring board was established before the start of the trials to provide oversight of the safety data during the studies. The protocols and informed consents were approved by the institutional review board of JPCDC. Written informed consent from all participants was obtained before screening for eligibility.

Eligible participants were healthy adults aged 18 through 59 years, who were seronegative for SARS-CoV-2 IgM and IgG and negative for SARS-CoV-2 nucleic acid as confirmed by pharyngeal swab reverse transcription polymerase chain reaction (RT-PCR). Confirmed cases, suspected cases or asymptomatic cases with COVID-19 as referred to the Information System of China Disease Prevention and Control are excluded. People having close contact with confirmed or suspected cases, travel history of abroad or domestic epidemic community within 14 days before vaccination are also excluded. To be included, participants should have an axillary temperature of 37·0°C or less; and have general good health as established by medical history, physical examination, and lab testing. Pregnant or breastfeeding women were excluded. People with previous SARS-CoV infection, mental disease, allergic reaction to any ingredient included in this vaccine or severe allergy to any other vaccines, congenital or acquired immune deficiency, HIV infection, serious systemic diseases, or other major chronic illnesses were also excluded. A complete list of the inclusion and exclusion criteria is provided in the protocol.

### Randomisation and masking

KCONVAC was developed by Shenzhen Kangtai Biological Products Co., Ltd. (Shenzhen, China) and Beijing Minhai Biotechnology Co., Ltd. (Beijing, China). The SARS-CoV-2 virus strain CQ01 was inoculated in Vero cell for cultivation. The harvested virus was inactivated by β-propiolactone, purified, and adsorbed to aluminum hydroxide (adjuvant). Each dose of vaccine contained 5 μg or 10 μg of inactivated SARS-CoV-2 virus antigen and 0·25 mg of aluminum in a 0·5-mL liquid formulation. The placebo contained the same adjuvant, with no virus antigen. The experimental vaccines and placebo were blindly labelled with a randomisation number on each vial as the only identifiers.

In the phase 1 trial, a randomisation ratio of 4:1 was used for 5-μg vaccine versus placebo, and for 10-μg vaccine versus placebo. In the phase 2 trial, the eligible participants were first stratified by vaccination regimen, then randomized in each stratum in a ratio of 2:2:1 to receive either 5-μg vaccine, 10-μg vaccine, or placebo. The randomisation list was generated by an independent statistician using SAS software (version 9·4). The participants received vaccine or placebo labelled with the same randomization number. Individuals involved in randomisation and masking had no involvement in the rest of the trial. Participants, investigators, and staff undertaking lab testing were masked to treatment allocation.

### Procedures

The phase 1 trial of KCONVAC was conducted in a manner of dosage escalation and prior to the initiation of the phase 2 trial. The first 30 eligible participants enrolled for the phase 1 trial were randomized to receive either vaccine of 5 μg or placebo. The safety profile within seven days post the first dose of 5-μg vaccine or placebo were assessed. If it was acceptable, the remained 30 participants for the phase 1 trial would be enrolled and randomized to receive either vaccine of 10 μg or placebo. The participants who received placebo in the phase 1 trial would be combined into one group for analysis. The safety profile within seven days post the first dose of 10-μg vaccine or placebo were assessed. If it was acceptable, the phase 2 trial would be initiated. One vaccination regimen was used in the phase 1 trial, i.e., two doses administered intramuscularly on Day 0 and Day 14 (0/14). Two vaccination regimens were used in the phase 2 trial, i.e., two doses administered intramuscularly on Day 0 and Day 14 (0/14), or Day 0 and Day 28 (0/28).

Participants were observed for 30 minutes following each vaccination for any immediate reaction, and received diary cards to record adverse events occurred within seven days after each vaccination. The adverse events occurred from Day 8 through Day 28 (when applicable) after each vaccination would also be recorded. To verify the adverse events, on-site visit would be done on Day 3, 7, 14, and 28 (when applicable) after each vaccination in the phase 1 trial, and on Day 7 and 28 (when applicable) after each vaccination in the phase 2 trial. Telephone contact would be done on Day 3 and 14 after each vaccination in the phase 2 trial. Blood biochemistry, hematology, blood coagulation function, and urinalysis were tested before vaccination and three days after each vaccination in the phase 1 trial. Adverse events were graded according to the scale issued by the National Medical Products Administration (NMPA), China in 2019.^16^

Blood samples for antibody assay were taken before vaccination, 14 and 28 days post the second dose from all the participants. Binding antibody responses against the receptor binding domain (RBD-IgG) of the spike glycoprotein of SARS-CoV-2 were tested by using ELISA with a detection limit of 1:20. Neutralising antibody responses were measured by using both live SARS-CoV-2 micro cytopathogenic effect assay with a detection limit of 1:4 and pseudovirus neutralisation tests (a vesicular stomatitis virus pseudovirus system expressing the spike glycoprotein) with a detection limit of 1:10.^17^

Undetectable antibody titre in serum was assigned a value of half the detection limits for calculation. For comparison of immune responses induced by natural SARS-CoV-2 infections, 35 convalescent serum samples were tested by micro cytopathogenic effect assay, which were collected 32-62 days after diagnosis by Hubei Provincial Center for Disease Control and Prevention. Antibody level of RBD-IgG subtypes including IgG1, IgG2, IgG3, IgG4 and antibody response to nucleoprotein of SARS-CoV-2 were determined by ELISA on Day 0, Day 14, Day 28 and Day 42. Cellular response was assayed in the phase 1 trial using ex-vivo interferon-γ (IFN-γ) enzyme-linked immunospot (ELISpot) on Day 0, Day 14, and Day 28, and serum cytokines test on Day 0, Day 14, Day 28, and Day 42. Positive IFNγ-ELISpot response was defined as the difference of average spot-forming cells per 200,000 peripheral blood mononuclear cells between stimulated and non-stimulated wells was greater than six, and the ratio was greater than two.

### Outcomes

In the phase 1 trial, the primary endpoint for safety was the proportion of participants experiencing adverse reactions/events within 28 days following each vaccination. The secondary endpoints included occurrence of serious adverse events (SAE) from the first dose through 12 months post the second dose, abnormal changes in laboratory tests within three days following each vaccination, and the seroconversion and titre of RBD-IgG and neutralization antibody 14 and 28 days after each vaccination, and 3, 6, and 12 months after the second dose.

In the phase 2 trial, the primary endpoint for immunogenicity was the seroconversion and titre of neutralization antibody, and the seroconversion of RBD-IgG 28 days after the second dose. The secondary endpoints included the proportion of participants experiencing adverse reactions/events within 28 days following each vaccination, occurrence of SAE from the first dose through 12 months post the second dose, titre of RBD-IgG 28 days after the second dose, and the seroconversion and titre of RBD-IgG and neutralization antibody 14 days, and 3, 6, and 12 months after the second dose.

Seroconversion was defined as antibody titre: 1) < 1:4, < 1:30, or < 1:20 before vaccination and≥ 1:4, ≥ 1:30, or≥ 1:20 post vaccination for neutralization antibody against live SARS-CoV-2, neutralization antibody against pseudovirus, or RBD-IgG, respectively; or 2) ≥1:4, ≥1:30, or≥1:20 before vaccination and 4-fold or more increase post vaccination for the corresponding antibodies.

### Statistical analysis

The sample size for the phase 1 trial was not determined on the basis of statistical power calculations but in line with the guidance issued by NMPA for phase 1 vaccine trial. The results of immunogenicity from the phase 1 trial was not available when designing the phase 2 trial. Therefore, the sample size for the phase 2 trial was determined based on the assumption that the seroconversion percentages for live neutralisation antibody in vaccine group and placebo group were 80% and 30%, respectively. A sample size of 100 in vaccine group versus 50 in placebo group would have sufficient power (more than 99%) to demonstrate the difference in the seroconversion percentages for live neutralisation antibody between groups when tested with a two-sided alpha value of 0·05.

The participants who received at least one vaccination were included in safety analysis. The number and proportion of participants experiencing adverse reactions or events in each group were presented. The immunogenicity analysis was done in per-protocol set consisting of participants who did not deviate from the eligibility criterion, received two doses, donated blood samples as scheduled, and had evaluable immunogenic data. Immunogenicity was expressed using seroconversion percentage, geometric mean titre (GMT), and the associated 95% confidence interval (CI). Antibody titre of individuals was log-transformed to calculate GMT in groups. The χ2 test or Fisher’s exact test was used to compare difference between groups for categorical data. ANOVA was used to test difference between groups for log-transformed antibody titres. The studies were registered with ClinicalTrials.gov, number NCT04758273 and NCT04756323.

## Results

The trial profile is shown in Figure 1. A total of 60 participants were enrolled in the phase 1 trial, received at least one dose of 5-μg vaccine (N=24), 10-μg vaccine (N=24), or placebo (N=12), and were included in safety analysis. One participant (5-μg vaccine group) discontinued the study and was not included in immunogenicity analysis. A total of 500 participants were enrolled in the phase 2 trial, received at least one dose of 5-μg vaccine (N=100 for each 0/14 or 0/28 regimen), 10-μg vaccine (N=100 for each regimen), or placebo (N=50 for each regimen), and were included in safety analysis. Five and four participants from 0/14 and 0/28 regimens respectively, discontinued the study and were not included in immunogenicity analysis. The average age across the treatment groups were 38·0 to 46·2 years of age. Baseline characteristics were generally similar between groups (Table 1). The two studies are ongoing to continuously follow up the safety and antibody persistence as planned. The preliminary analysis presents the data through the cutoff of 28 days post the second vaccination.

**Table 1.** Baseline characteristics of the study participants.

|  | Phase 1 trial (0/14) |  |  | Phase 2 trial (0/14) |  |  | Phase 2 trial (0/28) |  |  |
| --- | --- | --- | --- | --- | --- | --- | --- | --- | --- |
|  | 5 µg (N=24) | 10 µg (N=24) | Placebo (N=12) | 5 µg (N=100) | 10 µg (N=100) | Placebo (N=50) | 5 µg (N=100) | 10 µg (N=100) | Placebo (N=50) |
| **Age (years), mean (SD)** | 38.0 (9.5) | 41.0 (10.3) | 38.3 (8.8) | 45.5 (9.3) | 44.9 (9.5) | 46.2 (9.2) | 42.4 (10.5) | 44.5 (10.7) | 41.7 (10.0) |
| **Sex, n (%)** |  |  |  |  |  |  |  |  |  |
| **Male** | 12 (50%) | 10 (42%) | 8 (67%) | 53 (53%) | 45 (45%) | 19 (38%) | 38 (38%) | 46 (46%) | 25 (50%) |
| **Female** | 12 (50%) | 14 (58%) | 4 (33%) | 47 (47%) | 55 (55%) | 31 (62%) | 62 (62%) | 54 (54%) | 25 (50%) |
| **Completed, n (%)** | 23 (96%) | 24 (100%) | 12 (100%) | 100 (100%) | 97 (97%) | 48 (96%) | 98 (98%) | 99 (99%) | 49 (98%) |
| **Discontinued, n (%)** | 1 (4%) | 0 | 0 | 0 | 3 (3%) | 2 (4%) | 2 (2%) | 1 (1%) | 1 (2%) |
| **Neutralising antibody to live SARS-CoV-2** |  |  |  |  |  |  |  |  |  |
| **Seropositive** | 0 | 0 | 0 | 0 | 0 | 0 | 0 | 0 | 0 |
| **GMT** | 2 (2-2) | 2 (2-2) | 2 (2-2) | 2 (2-2) | 2 (2-2) | 2 (2-2) | 2 (2-2) | 2 (2-2) | 2 (2-2) |
| **Neutralising antibody to pseudovirus** |  |  |  |  |  |  |  |  |  |
| **Seropositive** | 0 | 0 | 0 | 6 (6%, 2-13) | 1 (1%, 0-5) | 0 | 2 (2%, 0-7) | 2 (2%, 0-7) | 2 (4%, 0-14) |
| **GMT** | 8 (6-10) | 10 (8-12) | 7 (5-9) | 8 (7-9) | 8 (7-9) | 7 (6-8) | 11 (9-12) | 9 (8-10) | 9 (8-10) |
| **RBD-IgG** |  |  |  |  |  |  |  |  |  |
| **Seropositive** | 0 | 0 | 0 | 0 | 0 | 0 | 0 | 0 | 1 (2%, 0-11) |
| **GMT** | 10 (10-10) | 10 (10-10) | 10 (10-10) | 11 (10-13) | 11 (10-12) | 10 (10-10) | 10 (10-12) | 10 (10-11) | 10 (10-11) |

Data are GMT (95% CI), number of participants (%; 95% CI) for seropositive (antibody titre ≥ detection limit). N=number of participants randomized in each treatment group. SARS-CoV-2=severe acute respiratory syndrome coronavirus 2. GMT=geometric mean titre. RBD-IgG= antibody to receptor binding domain; 0/14 (or 0/28): participants received two doses on Day 0 and Day 14 (or Day 0 and Day 28).

**Figure 1:**
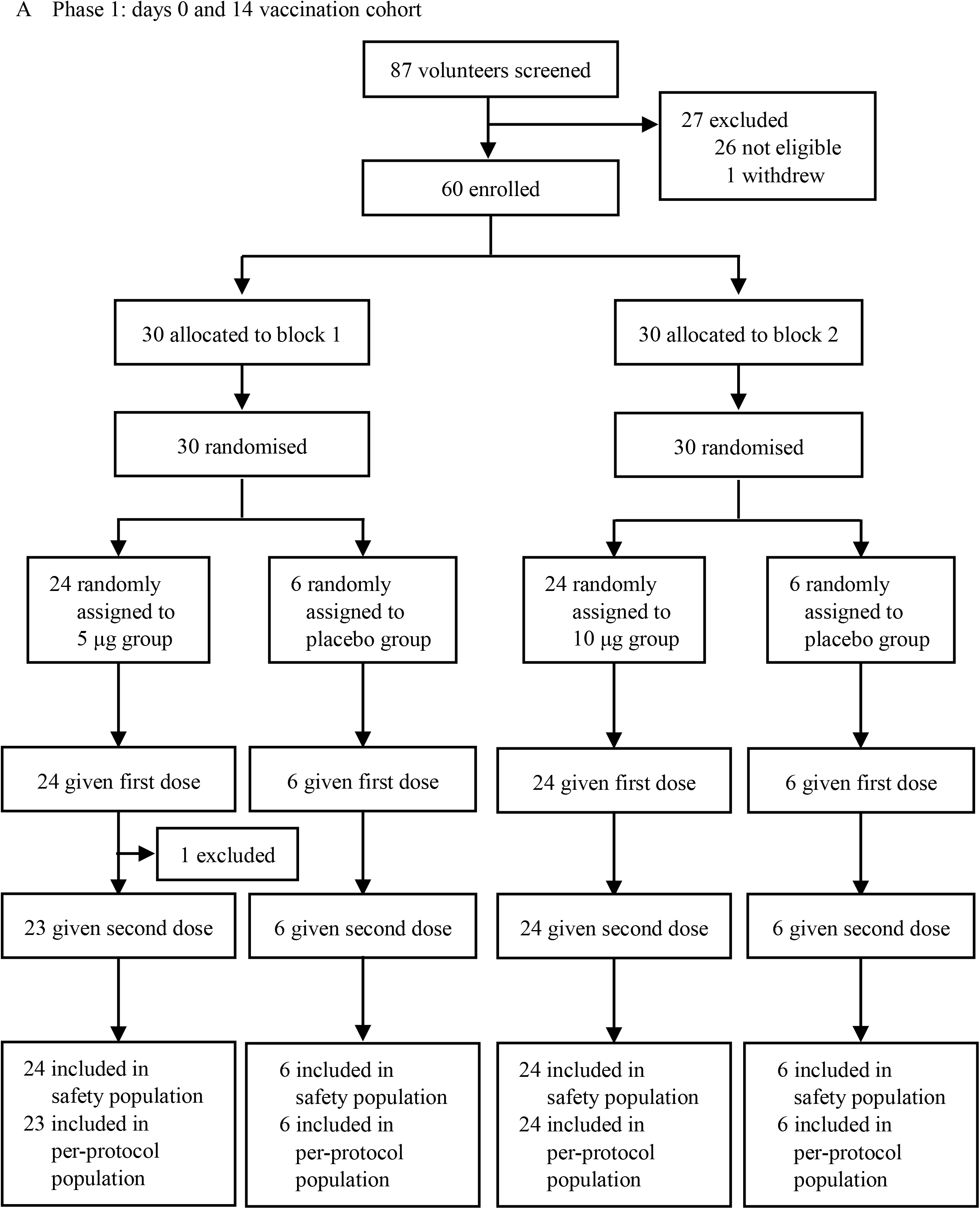
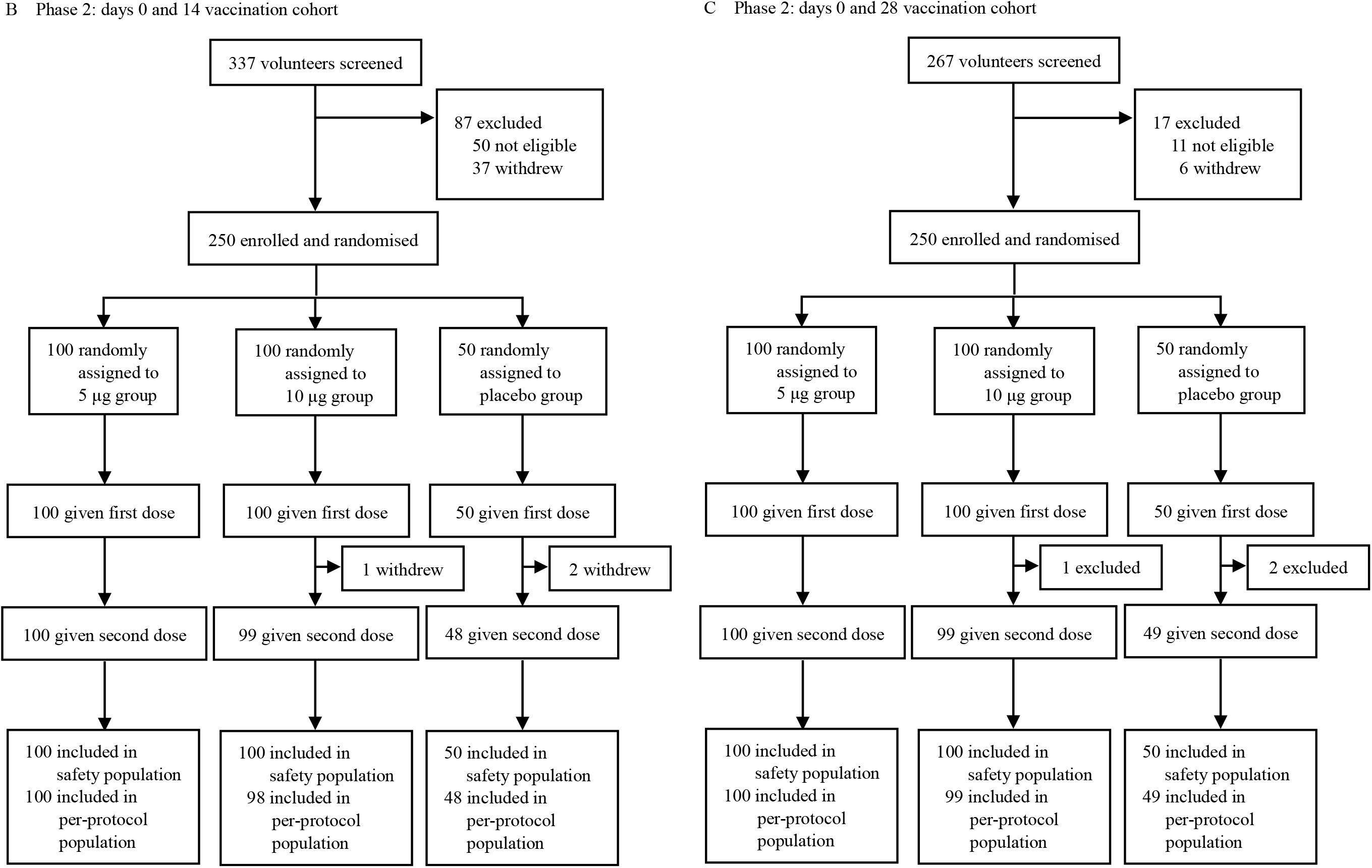
Study profile.

In the phase 1 trial, 13 (54%), 11(46%), and 7 (58%) participants reported at least one AE, of whom 10 (42%), 6 (25%), and 6 (50%) participants reported at least one vaccination-related AE after receiving 5-μg vaccine, 10-μg vaccine, or placebo, respectively. All the AEs were grade 1 or 2 in intensity. No AE of grade 3 or more was reported. The most common solicited injection-site AE and systemic AE across the three treatment groups were pain and fatigue, respectively (Table 2). One SAE (foot fracture) was reported in 10-μg vaccine group. No participant discontinued the study due to AE. 14 participants experiencing vaccination-related abnormal changes in blood biochemistry, hematology, or urinalysis across the three treatment groups with no statistical difference between vaccine groups and placebo group except hemobilirubin elevated (Supplementary Table S1).

**Table 2.** Adverse events within 28 days following vaccination.

|  | Phase 1 trial (0/14) |  |  | Phase 2 trial (0/14) |  |  | Phase 2 trial (0/28) |  |  |
| --- | --- | --- | --- | --- | --- | --- | --- | --- | --- |
|  | 5 µg (N=24) | 10 µg (N=24) | Placebo (N=12) | 5 µg (N=100) | 10 µg (N=100) | Placebo (N=50) | 5 µg (N=100) | 10 µg (N=100) | Placebo (N=50) |
| **Any AE** | 13 (54%) | 11 (46%) | 7 (58%) | 16 (16%) | 19 (19%) | 9 (18%) | 25 (25%) | 26 (26%) | 11 (22%) |
| **Grade 3 or more** | 0 | 0 | 0 | 0 | 0 | 0 | 0 | 0 | 0 |
| **Vaccination-related AE** | 10 (42%) | 6 (25%) | 6 (50%) | 13 (13%) | 17 (17%) | 6 (12%) | 19 (19%) | 24 (24%) | 9 (18%) |
| **Solicited Injection-site AE** | 4 (17%) | 4 (17%) | 2 (17%) | 11 (11%) | 8 (8%) | 4 (8%) | 16 (16%) | 18 (18%) | 7 (14%) |
| **Induration** | 1 (4%) | 0 | 0 | 2 (2%) | 0 | 0 | 0 | 3 (3%) | 0 |
| **Swelling** | 0 | 0 | 0 | 0 | 0 | 0 | 0 | 2 (2%) | 0 |
| **Erythema** | 2 (8%) | 0 | 0 | 1 (1%) | 0 | 0 | 1 (1%) | 5 (5%) | 0 |
| **Pain** | 3 (13%) | 4 (17%) | 2 (17%) | 8 (8%) | 8 (8%) | 4 (8%) | 15 (15%) | 12 (12%) | 7 (14%) |
| **Pruritus** | 0 | 0 | 0 | 2 (2%) | 1 (1%) | 1 (2%) | 1 (1%) | 2 (2%) | 0 |
| **Solicited systemic AE** | 2 (8%) | 1 (4%) | 1 (8%) | 6 (6%) | 11 (11%) | 4 (8%) | 6 (6%) | 10 (10%) | 2 (4%) |
| **Fever** | 0 | 0 | 0 | 1 (1%) | 2 (2%) | 1 (2%) | 1 (1%) | 1 (1%) | 1 (2%) |
| **Diarrhea** | 0 | 0 | 0 | 0 | 2 (2%) | 1 (2%) | 1 (1%) | 2 (2%) | 0 |
| **Inappetence** | 0 | 0 | 0 | 1 (1%) | 1 (1%) | 0 | 0 | 0 | 0 |
| **Vomiting** | 0 | 0 | 0 | 0 | 1 (1%) | 0 | 0 | 0 | 0 |
| **Nausea** | 0 | 0 | 0 | 0 | 0 | 0 | 0 | 1 (1%) | 0 |
| **Myalgia** | 1 (4%) | 0 | 0 | 1 (1%) | 1 (1%) | 1 (2%) | 1 (1%) | 0 | 0 |
| **Headache** | 0 | 0 | 0 | 2 (2%) | 6 (6%) | 0 | 2 (2%) | 0 | 1 (2%) |
| **Cough** | 0 | 0 | 1 (8%) | 0 | 3 (3%) | 0 | 1 (1%) | 3 (3%) | 0 |
| **Dyspnea** | 0 | 0 | 0 | 0 | 1 (1%) | 1 (2%) | 0 | 0 | 0 |
| **Skin or mucosa abnormality** | 0 | 0 | 0 | 0 | 1 (1%) | 0 | 0 | 0 | 0 |
| **Fatigue** | 2 (8%) | 1 (4%) | 1 (8%) | 2 (2%) | 6 (6%) | 0 | 2 (2%) | 3 (3%) | 1 (2%) |
| **Unsolicited AE** | 7 (29%) | 3 (13%) | 4 (33%) | 0 | 0 | 0 | 0 | 0 | 0 |

Data are n (%) of participants experiencing the relevant adverse events. N=number of participants included in each treatment group for the safety analysis. 0/14 (or 0/28): participants received two doses on Day 0 and Day 14 (or Day 0 and Day 28).

In the phase 2 trial, 16 (16%), 19 (19%), and 9 (18%) participants reported at least one AE, of whom 13 (13%), 17 (17%), and 6 (12%) participants reported at least one vaccination-related AE after receiving 5-μg vaccine, 10-μg vaccine, or placebo at the regimen of Day 0/14, respectively. Similar results were observed in the three treatment groups of Day 0/28 regimen. All the AEs were grade 1 or 2 in intensity. No AE of grade 3 or more was reported. The most common solicited injection-site AE and systemic AE across the six treatment groups were pain and fatigue, respectively (Table 2). No SAE was reported. No participant discontinued the study due to AE.

The baseline serostatus is summarized in Table 1. Before vaccination, the titres for neutralising antibody to live virus, neutralising antibody to pseudovirus, and RBD-IgG were quite low. Almost all participants were seronegative (under the detection limit) for the three antibodies. The vaccine induced significant antibody response (Table 3). After vaccinated with two doses, 87·5% (21/24) to 100% (24/24) of participants across the treatment groups in the phase 1 trial seroconverted for neutralising antibody to live virus, neutralising antibody to pseudovirus, and RBD-IgG 14 or 28 days post the second dose. Similar robust neutralising and RBD antibody responses were observed in the phase 2 trial where the vaccine induced seroconversion percentages of 83·0% (83/100) to 100% (99/99) across the treatment groups 14 or 28 days post the second dose. In contrast, in placebo group no (0/12) participant seroconverted for the three antibodies in the phase 1 trial, and only two (2/48) participants at Day 0/14 regimen and one (1/49) participant at Day 0/28 regimen seroconverted in the phase 2 trial. The differences in seroconversion percentages between vaccine groups and placebo groups are statistically significant (p<0.0001) for both dosages and both regimens, in both phase 1 and phase 2 trials.

**Table 3.** Antibody response 14 and 28 days post the second vaccination.

|  | 14 days post the second vaccination |  |  | 28 days post the second vaccination |  |  |
| --- | --- | --- | --- | --- | --- | --- |
|  | 5 µg | 10 µg | Placebo | 5 µg | 10 µg | Placebo |
| **Phase 1 trial (0/14)** |  |  |  |  |  |  |
| **N** | 23 | 24 | 12 | 23 | 24 | 12 |
| **Neutralising antibody to live SARS-CoV-2** |  |  |  |  |  |  |
| **Seroconversion** | 23 (100%, 85.2-100) | 23 (95.8%, 78.9-100) | 0 | 23 (100%, 85.2-100) | 24 (100%, 85.8-100) | 0 |
| **GMT** | 30.9 (20.6-46.4) | 40.6 (23.0-71.8) | 2.0 (2.0-2.0) | 29.3 (19.6-43.8) | 49.1 (33.5-72.0) | 2.0 (2.0-2.0) |
| **Neutralising antibody to pseudovirus** |  |  |  |  |  |  |
| **Seroconversion** | 22 (95.7%, 78.1-99.9) | 21 (87.5%, 67.6-97.3) | 0 | 22 (95.7%, 78.1-99.9) | 21 (87.5%, 67.6-97.3) | 0 |
| **GMT** | 90.4 (67.3-121.5) | 116.7 (71.1-191.6) | 7.8 (5.5-11.1) | 69.4 (53.8-89.6) | 99.8 (61.7-161.4) | 7.9 (5.4-11.4) |
| **RBD-IgG** |  |  |  |  |  |  |
| **Seroconversion** | 23 (100%, 85.2-100) | 24 (100%, 85.8-100) | 0 | 23 (100%, 85.2-100) | 24 (100%, 85.8-100) | 0 |
| **GMT** | 616.0 (381.9-993.7) | 1169.8 (694.2-1971.1) | 10.0 (10.0-10.0) | 605.3 (436.4-839.7) | 962.0 (613.5-1508.6) | 10.0 (10.0-10.0) |
| **Phase 2 trial (0/14)** |  |  |  |  |  |  |
| **N** | 100 | 98 | 48 | 100 | 97 | 48 |
| **Neutralising antibody to live SARS-CoV-2** |  |  |  |  |  |  |
| **Seroconversion** | 96 (96.0%, 90.1-98.9) | 95 (96.9%, 91.3-99.4) | 0 | 98 (98.0%, 93.0-99.8) | 96 (99.0%, 94.4-99.8) | 0 |
| **GMT** | 41.5 (32.8-52.5) | 48.3 (37.5-62.1) | 2.0 (2.0-2.0) | 37.2 (29.5-46.9) | 44.5 (35.5-55.7) | 2.0 (2.0-2.0) |
| **Neutralising antibody to pseudovirus** |  |  |  |  |  |  |
| **Seroconversion** | 87 (87.0%, 78.8-92.9) | 90 (91.8%, 84.6-96.4) | 0 | 83 (83.0%, 74.2-89.8) | 88 (90.7%, 83.1-95.7) | 0 |
| **GMT** | 94.4 (78.0-114.3) | 118.7 (96.4-146.2) | 7.7 (6.5-9.1) | 74.4 (62.6-88.5) | 97.6 (79.7-119.4) | 8.9 (7.5-10.6) |
| **RBD-IgG** |  |  |  |  |  |  |
| **Seroconversion** | 97 (97.0%, 91.5-99.4) | 95 (96.9%, 91.3-99.4) | 2 (4.2%, 0.5-14.3) | 98 (98.0%, 93.0-99.8) | 97 (100%, 96.3-100) | 1 (2.1%, 0.1-11.1) |
| **GMT** | 636.9 (496.5-816.7) | 652.7 (519.2-820.6) | 11.1 (9.4-13.0) | 623.0 (511.7-758.6) | 686.4 (574.2-820.1) | 10.5 (9.5-11.5) |

| Phase 2 trial (0/28) |  |  |  |  |  |  |
| --- | --- | --- | --- | --- | --- | --- |
| N | 100 | 99 | 49 | 98 | 99 | 49 |
| **Neutralising antibody to live SARS-CoV-2** |  |  |  |  |  |  |
| **Seroconversion** | 98 (98.0%, 93.0-99.8) | 99(100%, 96.3-100) | 0 | 97 (99.0%, 94.5-100) | 99 (100%, 96.3-100) | 0 |
| **GMT** | 110.5 (92.4-132.2) | 100.2 (84.6-118.7) | 2.0 (2.0-2.0) | 131.7 (109.3-158.6) | 110.7 (94.7-129.4) | 2.0 (2.0-2.0) |
| **Neutralising antibody to pseudovirus** |  |  |  |  |  |  |
| **Seroconversion** | 99 (99.0%, 94.6-100) | 98 (99.0%, 94.5-100) | 0 | 95 (96.9%, 91.3-99.4) | 96 (97.0%, 91.4-99.4) | 1 (2.0%, 0.1-10.9) |
| **GMT** | 276.6 (236.2-323.9) | 240.1 (204.3-282.2) | 8.1 (6.9-9.6) | 167.4 (142.6-196.5) | 153.6 (131.2-179.8) | 8.6 (7.2-10.2) |
| **RBD-IgG** |  |  |  |  |  |  |
| **Seroconversion** | 98 (98.0%, 93.0-100) | 98 (99.0%, 94.5-100) | 0 | 96 (98.0%, 92.8-99.8) | 99 (100%, 96.3-100) | 0 |
| **GMT** | 2485.5 (2051.2-3011.9) | 2037.3 (1643.4-2525.5) | 10.2 (9.8-10.6) | 1594.0 (1334.4-1904.1) | 1496.8 (1255.9-1783.9) | 10.3 (9.7-10.8) |

Data are GMT (95% CI), number of participants (% , 95% CI) for seroconversion. N=number of participants included in each treatment group for the per-protocol immunogenicity analysis.
SARS-CoV-2=severe acute respiratory syndrome coronavirus 2. GMT=geometric mean titre. RBD-IgG= antibody to receptor binding domain; 0/14 (or 0/28): participants received two doses on Day 0 and Day 14 (or Day 0 and Day 28).

Antibody titres rose to a high level after two-dose vaccination. Across the treatment groups in the two trials, the GMTs of neutralising antibody to live virus ranged from 29·3 to 49·1 at Day 0/14 regimen and from 100·2 to 131·7 at Day 0/28 regimen, neutralising antibody to pseudovirus ranged from 69·4 to 118·7 at Day 0/14 regimen and from 153·6 to 276·6 at Day 0/28 regimen, and RBD-IgG ranged from 605·3 to 1169·8 at Day 0/14 regimen and from 1496·8 to 2485·5 at Day 0/28 regimen, which were significantly elevated from the baseline titres. Correlation coefficients are 0.65 between live neutralization antibody and pseudovirus neutralization antibody, 0.66 between live neutralization antibody and RBD-IgG, and 0.69 between pseudovirus neutralization antibody and RBD-IgG. The GMT of neutralising antibody to live virus observed in convalescent serum was 49·7 (95%CI 33·3-74·3) (Figure 2).

**Figure 2:**
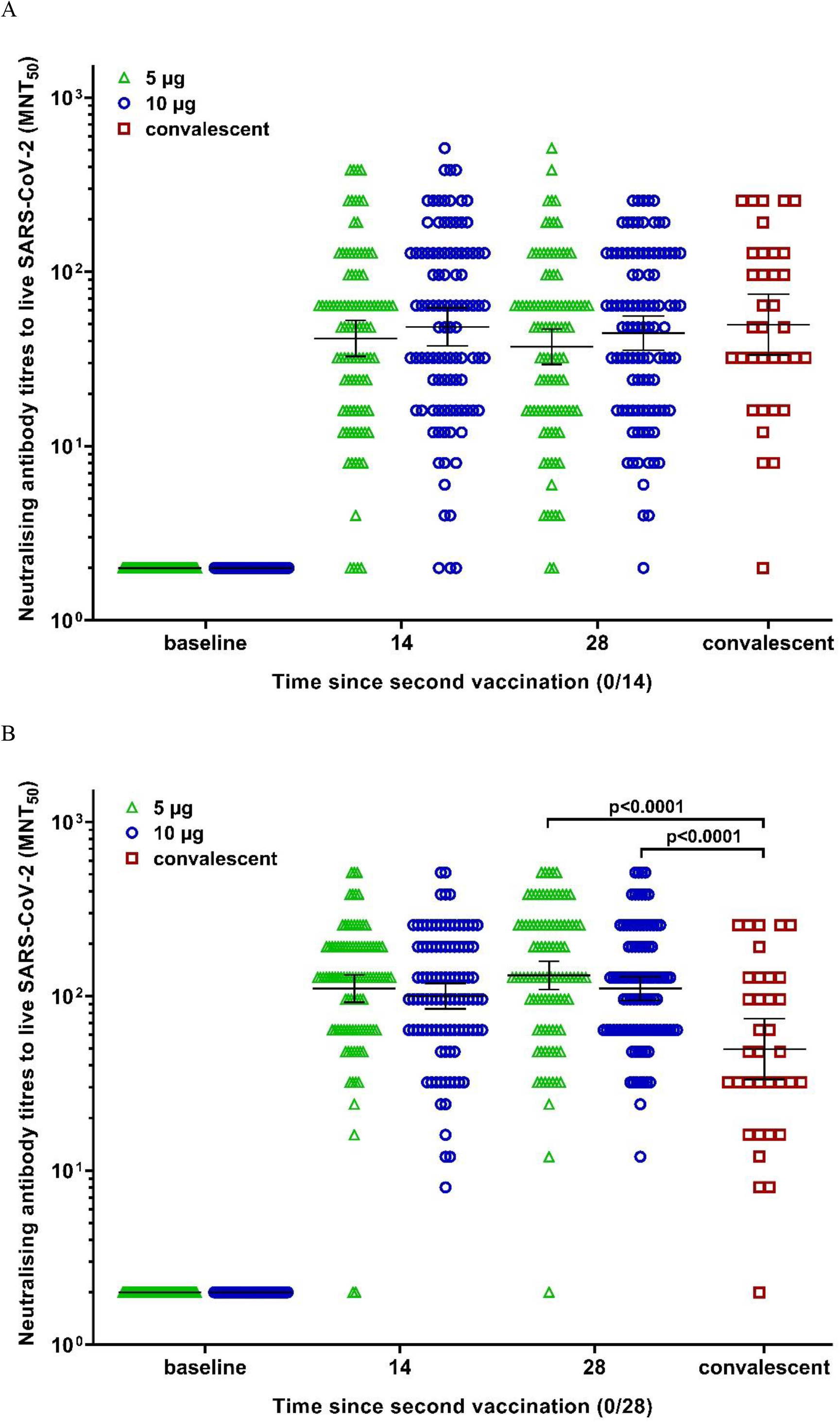
Neutralising antibody titer to live SARS-CoV-2 in the phase 2 trial and convalescent sera. 0/14=the regimen of Day 0/14 (A). 0/28=the regimen of Day 0/28 (B). The horizontal bars show the GMTs, the error bars indicate the 95% CIs of the GMTs and the dots indicated the individual antibody titers.

RBD-IgG subtyping assay showed that a significant part of RBD-IgG was IgG1, and a small part was IgG4. IgG2 and IgG3 were almost not detected. The RBD-IgG subtype GMT ratios (IgG1/IgG4) were 2.8 and 2.5 in 5-μg vaccine and 10-μg vaccine group 14 days after the second dose, respectively. In addition, a high level of anti-nucleocapsid protein antibody (N-IgG) was detected with GMT from 122·0 to 394·7 in vaccine groups 28 days after the second dose (Table 4).

**Table 4.** Subtyping assay for RBD-IgG and titration for N-IgG in the phase 1 trial.

|  | baseline |  |  | Day 28 |  |  | Day 42 |  |  |
| --- | --- | --- | --- | --- | --- | --- | --- | --- | --- |
|  | 5 µg (N=24) | 10 µg (N=24) | Placebo (N=12) | 5 µg (N=23) | 10 µg (N=24) | Placebo (N=12) | 5 µg (N=23) | 10 µg (N=23) | Placebo (N=12) |
| **IgG1 GMT** | 5.0 (5.0-5.0) | 5.0 (5.0-5.0) | 5.0 (5.0-5.0) | 34.4 (25.2-47.0) | 42.4 (30.0-59.8) | 5.0 (5.0-5.0) | 30.5 (25.6-37.1) | 31.4 (22.5-43.9) | 5.0 (5.0-5.0) |
| **IgG2 GMT** | 5.0 (5.0-5.0) | 5.0 (5.0-5.0) | 5.0 (5.0-5.0) | 5.2 (4.8-5.5) | 5.5 (4.6-6.5) | 5.0 (5.0-5.0) | 5.0 (5.0-5.0) | 5.3 (4.7-6.0) | 5.0 (5.0-5.0) |
| **IgG3 GMT** | 5.0 (5.0-5.0) | 5.0 (5.0-5.0) | 5.0 (5.0-5.0) | 5.8 (4.9-7.0) | 6.7 (5.2-8.5) | 5.0 (5.0-5.0) | 5.2 (4.8-5.5) | 5.8 (4.8-7.1) | 5.0 (5.0-5.0) |
| **IgG4 GMT** | 5.0 (5.0-5.0) | 5.0 (5.0-5.0) | 5.0 (5.0-5.0) | 12.0 (9.0-16.0) | 17.3 (12.1-24.7) | 5.0 (5.0-5.0) | 9.4 (7.0-12.7) | 12.7 (9.2-17.5) | 5.0 (5.0-5.0) |
| **N-IgG GMT** | 11.9 (7.2-19.7) | 10.0 (6.7-14.9) | 6.3 (4.7-8.4) | 90.3 (50.4-161.7) | 358.8 (208.9-616.2) | 5.6 (4.7-6.7) | 122.0 (72.2-206.2) | 394.7 (240.6-647.8) | 6.3 (4.5-8.9) |

Data are GMT (95% CI). GMT=geometric mean titre. RBD-IgG= antibody to receptor binding domain; N-IgG=antibody to nucleoprotein. N=number of participants included in each treatment group for the per-protocol immunogenicity analysis.

The vaccine induced obvious T-cell response with 56·5% (13/23) and 62·5% (15/24) of participants in 5-μg and 10-μg vaccine groups showed positive IFNγ-ELISpot responses 14 days after the second dose in the phase 1 trial. The IFN-γ positive SFCs per 200000 cells were 14·8 and 24·3 in the two vaccine groups. In contrast, IFNγ-ELISpot response was not detected in placebo group (Figure 3 and Supplementary Table S2). Serum IL-2 was detected in a significant part of participants 14 or 28 days after vaccination. However, other serum cytokines were not or less detected (Supplementary Table S3).

**Figure 3:**
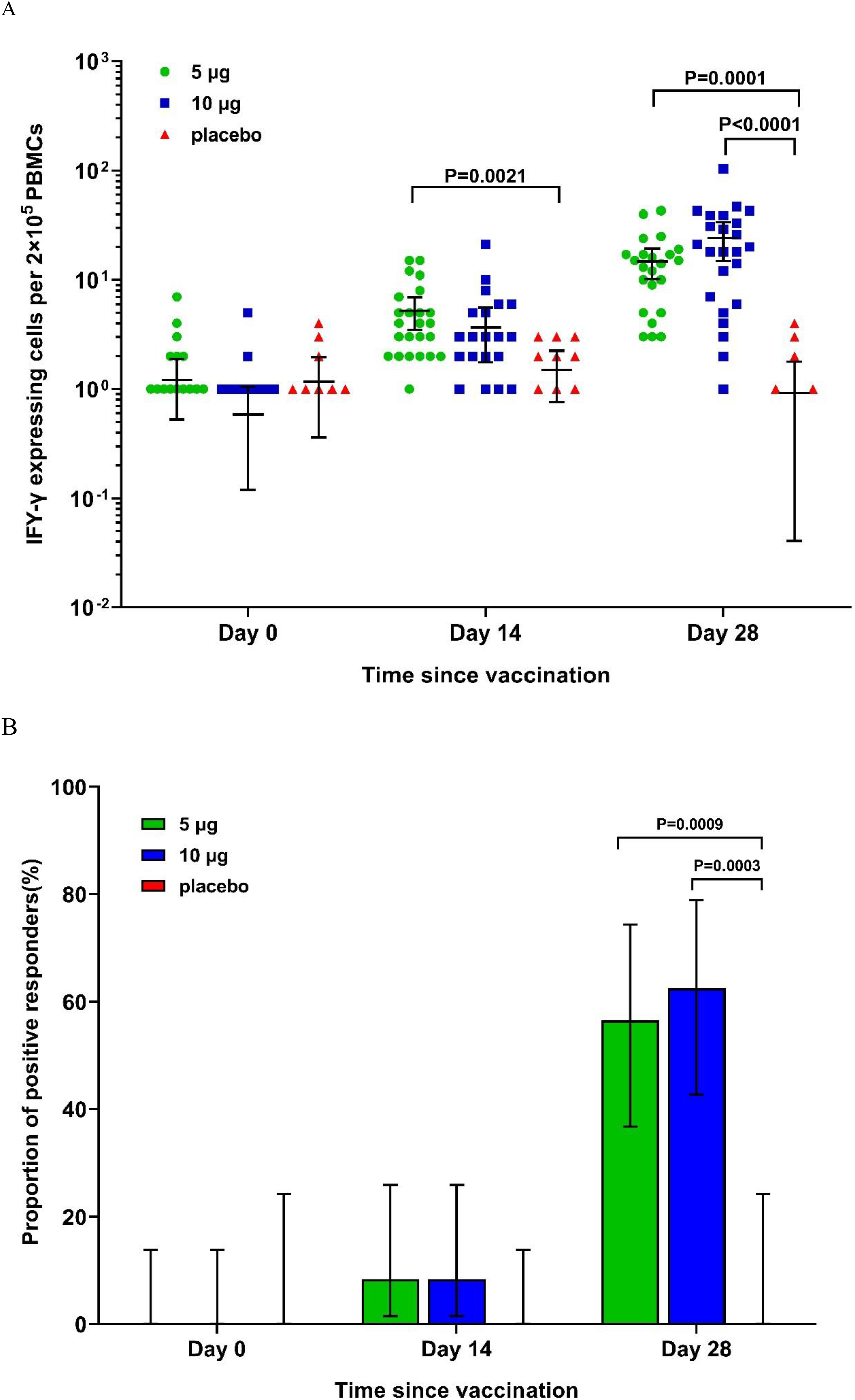
Specific T-cell responses measured by ELISpot in the phase 1 trial. IFN-γ positive SFCs per 200000 cells (A); Proportion of participants showing positive IFNγ-ELISpot response (B). IFN=interferon. PBMC=peripheral blood mononuclear cell.

## Discussion

Taking into account the urgent needs for vaccines against COVID-19 and the well-documented safety record of various inactivated vaccines, we conducted the phase 1 and phase 2 trials of KCONVAC in succession to accelerate the clinical development. When the preliminary safety data seven days following the first dose of 5-μg vaccine in the phase 1 trial were assessed and deemed acceptable, the administration of 10-μg vaccine in the phase 1 trial would start. The same requirement was applied when going forward from 10-μg vaccine group of the phase 1 trial to phase 2 trial. This intended to ensure the studies were conducted in a dosage-escalation and scale-up manner to safeguard the safety of the participants, and at the same time to accelerate the clinical process.

The preliminary safety analysis using the data following the first dose through 28 days post the second dose showed that KCONVAC is well tolerated in the study population. Proportions of participants experiencing AE or vaccination-related AE among the three treatment groups of the phase 1 trial are quite similar (p=0·31 or 0·84, respectively). In the phase 2 trial, the six treatment groups did not show significant difference regarding the proportions of participants experiencing AE or vaccination-related AE (p=0·61 to 0·91). The injection-site AEs and systemic AEs observed in our trials are common with other vaccines used in routine immunization practice. The safety profile of this vaccine is similar to other inactivated SARS-CoV-2 vaccines.^7,8^ No severe (grade 3 or more) AE was observed in the two trials.

The GMTs of the three antibodies in the phase 2 trial were generally comparable between the two timepoints, i.e., 14 or 28 days post the second dose, except that GMTs of neutralising antibody to pseudovirus were higher 14 days post the second dose than 28 days post the second dose for both 5-μg vaccine (p<0·0001) and 10-μg vaccine (p=0·0001), and that GMT of RBD-IgG was higher 14 days post the second dose than 28 days post the second dose for 5-μg vaccine (p=0·0009), when administered at the regimen of Day 0/28. This suggests that the antibody response might mount a peak between Day 14 and Day 28 post the second dose. Antibody dynamics needs more study. Our ongoing study will continue monitor the antibody persistence.

In the phase 2 trial, the regimen of Day 0/28 induced higher GMT than the regimen of Day 0/14 did, for both 5-μg vaccine and 10-μg vaccine. 14 days post the second dose, the GMTs induced by Day 0/28 regimen are 2-to-3·9 times of the GMTs induced by Day 0/14 regimen (p<0·0001 for all comparisons), and 28 days post the second dose, the GMTs induced by Day 0/28 regimen are 1·6-to-3·5 times of the GMTs induced by Day 0/14 regimen (p≥ 0·0005 for all comparisons). The observation that longer interval between dosing may induce higher antibody titre is consistent with another inactivated SARS-CoV-2 vaccine and other inactivated vaccines such as inactivated polio vaccine.^7,18^ The GMT of neutralising antibody to live virus of human convalescent serum was not significantly different comparing with that of the regimen of Day 0/14 (p≥ 0·35 for all comparisons), but significantly lower than that of the regimen of Day 0/28 (p<0·0001 for all comparisons), both in 5-μg vaccine and 10-μg vaccine.

Dosage-dependent antibody response was observed in the phase 1 trial and phase 2 trial (Day 0/14 regimen) at the two time points, i.e., 14 or 28 days post the second dose, in which 10-μg vaccine intended to induce higher GMTs across the three antibodies than 5-μg vaccine did (up to 1·9 fold). However, it was not observed in the phase 2 trial (Day 0/28 regimen). The exact reason is not known. As observed in the phase 2 trial, longer interval between dosing may induce higher antibody titre. The impact of longer interval might mitigate to some extent the effect of higher dosage.

Numerically, GMT of RBD-IgG was higher than that of neutralising antibodies to both live virus and pseudovirus, and GMT of neutralising antibody to pseudovirus was higher than that to live virus when vaccines were given at the same regimen with the same dosage and antibody titres were assayed at the same time points, which was consistently observed in previous studies. ^5 6^ Correlation analysis shows that the three antibodies are less correlated with lower coefficients than previous reports. More studies are needed to explore the correlation among various antibodies to optimize antibody assay methodology.

Antibody response to proteins generally induce primarily IgG1, accompanying with low level of IgG3 and IgG4, and IgG3 has even a short seven-days half-life^19^. This might explain why IgG2 and IgG3 were almost not detected. In humans, Th1 cells are considered to be associated with generation of IgG1 and IgG3, while Th2 cells are associated with generation of IgG4. ^20^ RBD-IgG1 GMT was approximately 3 times of RBD-IgG4 GMT on Day 28 and Day 42, which might indicate a Th1-biased response. The results of IFNγ assayed by ELISpot might be the evidence of the Th1-biased response. In addition, IgG1 has a longer half-time, indicating a favor to antibody persistence. Serum cytokines assay detected IL-2 but not other cytokines in a significant proportion of participants. It is well known that cytokines play a role in inflammation. Therefore, the cytokines profile observed 14 or 28 days after dosing might have relevance to adverse events, indicating that adverse events might be transient and disappear soon.

One limitation for this preliminary analysis is that we don’t include the safety and immunogenicity data of KCONVAC in elder adults who are at higher risk to COVID-19. These population are included in our protocol and will be the target in our ongoing studies. The second limitation is that the study duration of the trials is relative short, i.e., 28 days post the second dose. Therefore, long-term safety profile and antibody persistence are not in the scope of this preliminary analysis. Our ongoing studies and the upcoming phase 3 efficacy trial will be able to include these objectives. The third limitation is that we used ELISA instead of ELISpot to test the cytokines other than IFNγ and collected blood samples only once between the first and second doses. Therefore we could not get an overall profile of the cell response. The reason for using ELISA and this sampling schedule was that we intended to observe the safety only through the serum cytokines, and considered that inactivated virus vaccine might show weak cellular response.

In conclusion, our two trials demonstrate that KCONVAC is well tolerated and able to induce robust antibody response and cellular response in adults aged 18 to 59 years, which warrants further evaluation with this vaccine in the upcoming phase 3 efficacy trial.

## Research in context

### Evidence before this study

We searched PubMed on March 13, 2021, for clinical trial reports published on peer-reviewed journals with the terms “COVID-19” or “SARS-CoV-2”, “vaccine”, and “clinical trial” or “trial”, and found 29 original articles reporting the safety, immunogenicity and/or efficacy of SARS-CoV-2 vaccines in human which were developed using the platforms or technologies of inactivated- (6), adenovirus vectored- (12), recombinant protein-(2), RNA- (8), and DNA-(1) based vaccines. In addition, using the same terms we also identified three relevant articles (one for inactivated vaccine, two for recombinant protein-based vaccine) published on medRxiv preprint. These vaccines demonstrated good immunogenicity and acceptable safety profile. Four vaccines also proved to be efficacious against symptomatic COVID-1. Some vaccines have been authorized for emergency use or received conditional approval from country regulatory authorities, including three inactivated vaccines and one adenovirus-vectored vaccine from China, two mRNA vaccines and one adenovirus-vectored vaccine from the United States, one adenovirus-vectored vaccine from the United Kingdom, and one adenovirus-vectored vaccine from Russia.

### Added value of this study

This study demonstrated good immunogenicity and tolerability of the experimental inactivated COVID-19 vaccine, KCONVAC, adding evidence that inactivated COVID-19 vaccines can induce both antibody response and cellular response, and that two doses spanning 14 or 28 days are needed to provoke robust immune response. Both neutralising antibody response and receptor binding domain antibody (RBD-IgG) response were elicited. The induced RBD-IgG was primarily IgG1, indicating a Th1-biased response. The good safety profile observed in this study continuously supports the development and deployment of inactivated vaccine for combating COVID-19.

### Implications of all the available evidence

Results from this study indicated that two doses of KCONVAC are of good immunogenicity and tolerability, warranting further evaluation in a phase 3 efficacy trial.

## Data Availability

Supporting clinical documents including study protocol and statistical analysis plan will be available immediately following publication for at least 1 year. Researchers who provide a scientifically sound proposal will be allowed access to the individual participant data. Proposals should be directed to. These proposals will be reviewed and approved by the funder, investigator, and collaborators on the basis of scientific merit. To gain access, data requesters will need to sign a data access agreement.

## Contributors

HP, BH, JianL, GL were co-first authors of this manuscript. WT, WH and FZ were joint corresponding authors. HP was the principal investigator of this trial. HP, JL, GL and FZ designed the trial and study protocol. YL contributed to the literature search. All authors had access to data and WT, WH and FZ verified the data. HP and JingL wrote the first draft the manuscript. WT, WH and FZ contributed to the data interpretation and revision of the manuscript. XC monitored the trial. KC, JH and HP were responsible for the site work including the recruitment, follow up, and data collection, and KC was the site coordinator. BH, WT and WH were responsible to the laboratory analysis.

## Acknowledgments

We appreciate the contributions of all investigators at the Jiangsu Provincial Center for Disease Control and Prevention (CDC) and Huaiyin CDC who worked on the trials. We thank Yuanzheng Qiu for helpful expert advice and support for writing. We would like to thank all the participants who volunteered for this study.

