## Supplementary materials for "Immunogenicity and Safety of a SARS-CoV-2 Inactivated Vaccine (KCONVAC) in Healthy Adults: Two Randomized, Double-blind, and Placebo-controlled Phase 1/2 Clinical Trials"

**Content**

### Table S1 Vaccination-related abnormal changes in blood biochemistry, hematology, or urinalysis

| **Abnormal change** | **5 μg (N=24)** | **10 μg (N=24)** | **Placebo (N=12)** | **P value** |
| --- | --- | --- | --- | --- |
| Urine protein positive | 2 (8%) | 1 (4%) | 1 (8%) | 1.0000 |
| Urine erythrocyte positive | 2 (8%) | 1 (4%) | 0 | 0.7980 |
| Hemobilirubin elevated | 0 | 0 | 3 (25%) | **0.0064** |
| Alanine aminotransferase elevated | 1 (4%) | 0 | 0 | 1.0000 |
| Lymphocytes count lowered | 1 (4%) | 0 | 0 | 1.0000 |
| Urine glucose positive | 0 | 1 (4%) | 0 | 1.0000 |
| Blood glucose elevated | 1 (4%) | 0 | 0 | 1.0000 |
| Sum | 7 (29%) | 3 (13%) | 4 (33%) | 0.2391 |

Data are n (%) of participants experiencing the relevant abnormal changes. N=number of participants included in each treatment group for the safety analysis.

### Table S2 Specific T-cell responses measured by ELISpot in the phase 1 trial

|  | **baseline** | | | **Day 14** | | | **Day 28** | | |
| --- | --- | --- | --- | --- | --- | --- | --- | --- | --- |
|  | **5 μg** | **10 μg** | **Placebo** | **5 μg** | **10 μg** | **Placebo** | **5 μg** | **10 μg** | **Placebo** |
| **N** | 24 | 24 | 12 | 24 | 24 | 12 | 23 | 24 | 12 |
| **Positive responders, n(%, 95%CI)** | 0 | 0 | 0 | 2 (8·3%, 1·0-27·0) | 2 (8·3%, 1·0-27·0) | 0 | 13 (56·5%, 34·5-76·8) | 15 (62·5%, 40·6-81·2) | 0 |
| **IFN-γ positive SFCs per 200000 cells** | 1·2 (0·5-1·9) | 0·9 (0·1-1·0) | 1·2 (0·4-2·0) | 5·2 (3·5-6·9) | 3·7 (1·8-5·6) | 1·5 (0·8-2·2) | 14·8 (10·2-19·3) | 24·3 (14·9-33·7) | 0·9 (0·0-1·8) |

### Table S3 Serum cytokines assay by ELISA in the phase 1 trial

|  | **baseline** | | | **Day 14** | | | **Day 28** | | | **Day 42** | | |
| --- | --- | --- | --- | --- | --- | --- | --- | --- | --- | --- | --- | --- |
|  | **5 μg** | **10 μg** | **Placebo** | **5 μg** | **10 μg** | **Placebo** | **5 μg** | **10 μg** | **Placebo** | **5 μg** | **10 μg** | **Placebo** |
| **TNF-α** |  |  |  |  |  |  |  |  |  |  |  |  |
| **N** | 24 | 24 | 12 | 24 | 24 | 12 | 23 | 24 | 12 | 23 | 24 | 12 |
| **Mean (SD)** | 1·4 (5·7) | 20·2 (33·7) | 13·1 (23·93) | 1·3 (5·5) | 0 | 0 | 1·5 (6·4) | 0 | 0 | 1·1 (4·9) | 0 | 0 |
| **Median** | 0 | 5·2 | 0 | 0 | 0 | 0 | 0 | 0 | 0 | 0 | 0 | 0 |
| **IL-2** |  |  |  |  |  |  |  |  |  |  |  |  |
| **N** | 24 | 24 | 12 | 24 | 24 | 12 | 23 | 24 | 12 | 23 | 24 | 12 |
| **Mean (SD)** | 0·1 (0·3) | 1·1 (4·2) | 0·5 (1·8) | 1·5 (4·7) | 0·7 (2·6) | 0·8 (2·6) | 3·7 (6·3) | 1·4 (5·1) | 2·2 (5·6) | 4·7 (7·0) | 3·2 (6·5) | 0 |
| **Median** | 0 | 0 | 0 | 0 | 0 | 0 | 0 | 0 | 0 | 0 | 0 | 0 |
| **IL-4** |  |  |  |  |  |  |  |  |  |  |  |  |
| **N** | 24 | 24 | 12 | 24 | 24 | 12 | 23 | 24 | 12 | 23 | 24 | 12 |
| **Mean (SD)** | 0 | 0 | 0 | 0 | 0 | 0 | 0·3 (1·1) | 0 | 0 | 0·1 (0·3) | 0·1 (0·5) | 0 |
| **Median** | 0 | 0 | 0 | 0 | 0 | 0 | 0 | 0 | 0 | 0 | 0 | 0 |
| **IL-5** |  |  |  |  |  |  |  |  |  |  |  |  |
| **N** | 24 | 24 | 12 | 24 | 24 | 12 | 23 | 24 | 12 | 23 | 22 | 12 |
| **Mean (SD)** | 0 | 0 | 0·8 (2·7) | 0 | 0 | 1·0 (3·5) | 0 | 0 | 1·1 (3·7) | 0 | 0·4 (1·7) | 0·5 (1·7) |
| **Median** | 0 | 0 | 0 | 0 | 0 | 0 | 0 | 0 | 0 | 0 | 0 | 0 |

Serum cytokines including TNF-α, IL-2, IL-4, IL-5,IL-6 and IFNγ was tested by ELISA on Day 0, 14, 28, 42. IL-6 and IFN-γ were not detected.
